# Infertility and pregnancy outcomes among adults with primary ciliary dyskinesia

**DOI:** 10.1101/2024.04.09.24305388

**Authors:** Leonie D Schreck, Eva S L Pedersen, Katie Dexter, Michele Manion, Living with PCD study advisory group, Nathalie Massin, Bernard Maitre, Myrofora Goutaki, Claudia E Kuehni

**Affiliations:** Institute of Social and Preventive Medicine, University of Bern, 3012 Bern, Switzerland; Graduate School for Health Sciences, University of Bern, 3012 Bern, Switzerland; PCD Support UK, Buckingham MK18 9DX, United Kingdom; PCD Foundation, Minneapolis, MN 55420, United States; IVF Center, American Hospital of Paris, 92200 Neuilly-sur-Seine, France; Université Paris Est Créteil, Institut national de la santé et de la recherche médicale (INSERM), IMRB, 94010 Créteil, France; Dept of Pneumology, Centre Hospitalier Intercommunal de Créteil, 94010 Créteil, France; Division of Paediatric Respiratory Medicine and Allergology, Department of Paediatrics, Inselspital, University Hospital, University of Bern, 3010 Bern, Switzerland

**Keywords:** primary ciliary dyskinesia, infertility, ectopic pregnancy, assisted reproduction, epidemiology

## Abstract

**Study question:** What is the prevalence of infertility and ectopic pregnancies among individuals with primary ciliary dyskinesia (PCD), to what extent do they benefit from medically assisted reproduction (MAR), and how does fertility differ by affected PCD gene?

**Summary answer:** We found that 39 of 50 men (78%) and 72 of 118 women (61%) with PCD were infertile. MAR was effective in infertile individuals, with around two-thirds of them successfully conceiving with MAR. Women with PCD had an increased risk of ectopic pregnancies (7.6 per 100 pregnancies, 95% CI 4.7-12.2). Our results suggest that fertility status in PCD differs by affected PCD gene.

**What is known already:** PCD is a heterogeneous multiorgan disease caused by mutations in genes required for the function and structure of motile cilia. Previous studies identified a link between PCD and infertility, but original data on prevalence of infertility and risk of ectopic pregnancies, the use and efficacy of MAR and the association of fertility with PCD genotype, are extremely limited.

**Study design, size, duration:** We performed a cross-sectional survey about fertility within the *Living with PCD* study (formerly COVID-PCD). *Living with PCD* is an international, online, participatory study that collects information directly from people with PCD. People with PCD of any age from anywhere in the world can participate in the study. At the time of the survey, 482 adults with PCD were registered within the *Living with PCD* study.

**Participants/materials, setting, methods:** We sent a questionnaire on fertility on July 12, 2022, to all participants older than 18 years enrolled in the *Living with PCD* study. The fertility questionnaire covered topics related to pregnancy attempts, use of MAR, and pregnancy outcomes. Data was collected via the Research Electronic Data Capture (REDCap) platform. We defined infertility as failure to achieve a clinical pregnancy after 12 months or use of MAR for at least one pregnancy (Zegers-Hochschild et al., 2017).

**Main results and the role of chance:** 265 of 482 adult participants (55%) completed the fertility questionnaire. Among 168 adults who had tried to conceive, 72 (61%) women and 39 (78%) men were infertile. Of the infertile men, 28 had tried MAR, and 17 of them (61%) fathered a child with the help of MAR. Among infertile women, 59 had used MAR, and 41 of them (69%) became pregnant with the help of MAR. In our population, women with PCD showed a relatively high risk of ectopic pregnancies: 1 in 10 women who became pregnant had at least one ectopic pregnancy and 7.6% of pregnancies were ectopic (95% CI 4.7-12.2). We evaluated the association between fertility and affected PCD genes in 46 individuals (11 men, 35 women) with available genetic and fertility information, and found differences between genotypes e.g. all 5 women with a mutation in CCDC40 were infertile and all 5 with DNAH11 were fertile.

**Limitations, reasons for caution:** The study has limitations, including potential selection bias as people experiencing problems with fertility might be more likely to fill in the questionnaire, which may have influenced our prevalence estimates. We were unable to validate clinical data obtained from participant self-reports due to the anonymous study design, which is likely to lead to recall bias.

**Wider implications of the findings:** The study underlines the need for addressing infertility in routine PCD care, with a focus on informing individuals with PCD about their increased risk. It emphasizes the utility and efficacy of MAR in PCD-related infertility. Additionally, women attempting conception should be made aware of the increased risk of ectopic pregnancies and seek systematic early consultation to confirm intrauterine pregnancy. Fertility, efficacy of MAR and risk for adverse pregnancy outcomes differ between people with PCD—depending on genotypes—, and close monitoring and support might be needed from fertility specialist to increase chances of successful conception.

**Study funding/competing interest(s):** Our research was funded by the Swiss National Science Foundation, Switzerland (SNSF 320030B_192804/1), the Swiss Lung Association, Switzerland (2021-08_Pedersen), and we also received support from the PCD Foundation, United States; the Verein Kartagener Syndrom und Primäre Ciliäre Dyskinesie, Germany; the PCD Support UK, United Kingdom; and PCD Australia, Australia. M. Goutaki received funding from the Swiss National Science Foundation, Switzerland (PZ00P3_185923). B. Maitre participates in the RaDiCo-DCP funded by INSERM France. Study authors participate in the BEAT-PCD Clinical Research Collaboration supported by the European Respiratory Society. All authors declare no conflict of interest.

**Trial registration number:** ClinicalTrials.gov ID NCT04602481

**What does this mean for patients?:** Primary ciliary dyskinesia (PCD) is a rare genetic disease. People who live with it can have problems conceiving. It is unclear how many people with PCD struggle to have children, and how many can only have children with help. We also do not know if women with PCD more often have ectopic pregnancies (= pregnancies outside of the uterus) than the general population. How did we answer these questions? We sent a questionnaire about fertility to all participants in the *Living with PCD* study. The *Living with PCD* study is an online study. It collects information directly from people with PCD from all over the world. How many people with PCD struggled to have children? Eight out of ten men and six out of ten women had problems conceiving. How many were successful with help?

Among those who struggled, two out of three were able to have a child with help of fertility treatments. Did women with PCD more often have ectopic pregnancies? Women with PCD more often had an ectopic pregnancy than the general population. In our study, seven out of 100 pregnancies were ectopic, compared to only two in 100 pregnancies in the general population. We believe that more people with fertility problems completed our questionnaire. Thus, the true risk of ectopic pregnancy in PCD might be lower than we found in our study. But ectopic pregnancies can lead to serious complications. Thus, the authors of this paper think that fertility specialists should inform women with PCD about their increased risk. Women with PCD should see their gynaecologist early in their pregnancy to confirm that the pregnancy is inside the uterus. The authors suggest to address fertility problems in routine PCD care, with the help of fertility specialists.

## Introduction

Primary ciliary dyskinesia (PCD) can lead to infertility and adverse pregnancy outcomes. PCD is a heterogeneous multiorgan disease that is caused by mutations in genes needed for the function and structure of motile cilia. More than 50 disease-causing genes have been described leading to varying degrees of PCD symptoms (Lucas et al., 2020, Wheway et al., 2021). Motile cilia lining the respiratory tract contribute to chronic airway disease, a common characteristic in patients with PCD. Furthermore, motile cilia play essential roles in the reproductive systems of men and women. In men with PCD, mutations in motile cilia in the efferent ductules may lead to oligozoospermia (reduced sperm count) or azoospermia (no sperm) due to obstructive sperm stasis (Aprea et al., 2021). Sperm flagella share similar axonemal structures with motile cilia, with mutations leading to asthenozoospermia (sperm dysmotility) or teratozoospermia (morphological abnormalities of sperm) (Sironen et al., 2020). In women, reduced function of motile cilia lining the fallopian tubes and the uterus may be the cause of reduced fertility (Lyons et al., 2006, Marchini et al., 1992, Pearson-Farr et al., 2023, Raidt et al., 2015), possibly leading to impaired oocyte transport and an increased risk of ectopic pregnancies (Blyth and Wellesley, 2008, Lin et al., 1998, McLean and Claman, 2000).

Counselling patients with PCD about their likelihood of conceiving is difficult and based on limited available data. We lack information about the extent of fertility issues and ectopic pregnancies and the use and success rates of medically assisted reproduction (MAR). Additionally, fertility is suggested to vary depending on the PCD gene affected but little is known about the exact associations. Evidence comes almost exclusively from case reports and case series. In a systematic review and meta-analysis from 2016, estimates of infertility prevalence were very heterogenous among 7 studies ranging from 15 to 79% (Goutaki et al., 2016). A recent narrative review confirmed the low level of evidence (Newman et al., 2023). The largest study on infertility to date from 2017 included 85 individuals from PCD hospitals in France and Belgium (Vanaken et al., 2017). 61% of women and 76% of men were considered infertile, and risk of infertility differed by involved PCD gene. In this study, 6 of 22 women and 15 of 37 men had benefitted from the use of MAR. Succes of MAR has also been described in case reports—particularly *in vitro* fertilization (IVF) for women and intracytoplasmic sperm injection (ICSI) for men (Jayasena and Sironen, 2021, Newman, et al., 2023). Data on the frequency of ectopic pregnancies is scarce with only 4 ectopic pregnancies reported in case reports so far (Blyth and Wellesley, 2008, Lin, et al., 1998, McLean and Claman, 2000, Newman, et al., 2023). It is impossible to extrapolate prevalence of ectopic pregnancies from these case reports.

In the largest study on fertility in PCD to date, we studied fertility and pregnancy outcomes of adults with PCD, including the prevalence of infertility, use and efficacy of MAR, the prevalence of ectopic pregnancies and the association of fertility with PCD genotype.

## Materials and methods

### Study design, study population, and ethics

We used data from a cross-sectional questionnaire about fertility conducted as part of the *Living with PCD* study (formerly COVID-PCD). *Living with PCD* is an international, online, participatory study that collects information directly from people with PCD. A detailed study protocol has been published (Pedersen et al., 2021). In short, *Living with PCD* was set up during the COVID-19 pandemic in 2020 at the University of Bern, Switzerland, in collaboration with PCD support groups worldwide. The study continuously monitored people with PCD during the pandemic and we have now started examining other aspects relevant to this population such as fertility. We identified this topic as of great interest by individuals living with PCD themselves based on the feedback from participants directly. Questionnaires are available in five languages (English, French, German, Italian, and Spanish) and people with PCD of any age from anywhere in the world can participate in the study. The study is advertised and distributed via PCD patient support groups worldwide and people with PCD or their caregivers, such as parents, actively contribute to study design. A study advisory group consisting of support group representatives is involved in all relevant decisions regarding the study. The study is anonymous, with participants or their caregivers providing informed consent upon study enrolment. Participants retain the right to withdraw from the study at any time by contacting the study team. The cantonal ethics committee of Bern, Switzerland, approved the study (study ID: 2020-00830).

### Study procedure

Participants self-registered via the study website (www.pcd.ispm.ch) and received a baseline questionnaire which included questions on demographic characteristics, diagnostic information including genetic results and symptoms, followed by shorter weekly follow-up questionnaires. We sent a special questionnaire dedicated to fertility on July 12, 2022, to all participants older than 18 years enrolled in the *Living with PCD* study. Non-responders received up to three reminders via email.

We collected responses until March 8, 2023. Participants entered data directly into an online database using the Research Electronic Data Capture (REDCap) platform (Harris et al., 2009) which is hosted by the Swiss medical registries and data linkage centre at the University of Bern, Switzerland.

### Fertility questionnaire

We developed the fertility questionnaire in close collaboration with fertility specialists, reproductive endocrinologists, and participants of the study advisory group where participants made suggestions to improve understanding of the questions. The questions and answer categories are in Supplementary Table S1. Members of the research team translated the questionnaires and two native speaking members of the team or from the study advisory group checked translations independently.

### Information on fertility and pregnancy

We asked participants if they had ever tried to conceive a child; if so, if they had used MAR, and if the attempt resulted in a pregnancy. We included all participants with available fertility data in our analyses and thus excluded participants who had never tried to conceive or had tried for less than 12 months (Figure 1). We categorised people as fertile if they achieved pregnancy naturally without MAR (Zegers-Hochschild, et al., 2017). Consequently, we defined them as infertile if they failed to achieve a clinical pregnancy after 12 months or if they used MAR for at least one pregnancy (World Health Organization (WHO), 2019/2021, Zegers-Hochschild, et al., 2017). We subclassified infertile individuals according to the use of MAR as “pregnancy with MAR”, “no pregnancy despite MAR”, “no pregnancy, no MAR”. We asked all participants about fertility tests and results, as well as other factors that may contribute to infertility, such as abnormal levels of fertility hormones or diseases of the reproductive system (Vander Borght and Wyns, 2018). The definition of these factors can be found in Supplementary Table S2.

**Figure 1:**
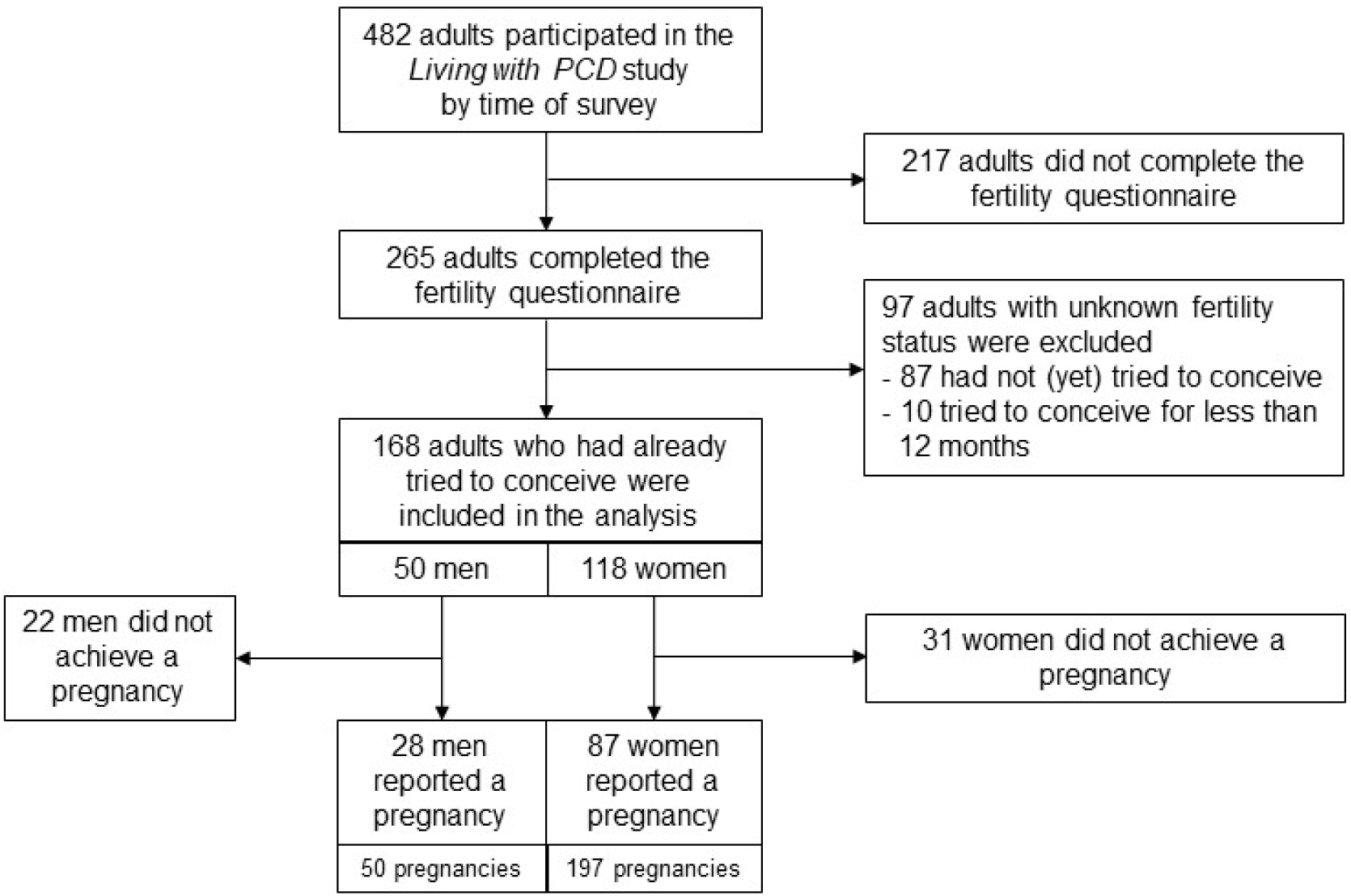
Flowchart of adults with primary ciliary dyskinesia in the *Living with PCD* study who were invited and completed the fertility questionnaire (N = 482).

Among participants reporting a pregnancy, we asked about the mode of fertilization and the outcome of the pregnancy. We grouped pregnancy outcome as live birth, pregnancy loss/stillbirth, ectopic pregnancy, abortion or ongoing pregnancy. To assess how fertility differed by genotype, we grouped reported PCD genes into categories based on associated defects: genes involved in the coding of proteins for dynein structure, dynein assembly, microtubular stabilization/nexin-dynein regulatory complex, radial spoke and central complex, and other function (Shoemark et al., 2021). We excluded participants who reported two heterozygous variants in different genes.

### Data analyses

We assessed the prevalence of infertility and fertility outcomes stratified by sex. We described use of MAR and examined the efficacy of MAR by determining the individuals who achieved pregnancy with MAR among all who underwent MAR. For women, we calculated the prevalence of ectopic pregnancies as the proportion of ectopic pregnancies among all reported pregnancies and all reported live births. Since not all *Living with PCD* participants completed the fertility questionnaire, to test the robustness of our findings, we also calculated the resulting minimum prevalence of ectopic pregnancies for our entire study population. We hypothesised that women with an ectopic pregnancy might have been more likely to fill in the questionnaire and assumed that women who did not complete the questionnaire had no ectopic pregnancy. For simplicity, we assumed that all women in *Living with PCD* had the same median number of pregnancies, regardless of questionnaire completion. Based on these two assumptions, we calculated the minimum prevalence as the proportion of reported ectopic pregnancies out of all hypothetical pregnancies and live births of all women in the *Living with PCD* study. Finally, we assessed fertility status by reported PCD genotype. We calculated Wilson 95% confidence intervals for proportions.

## Results

### Characteristics of the study participants

At the time of the study, 482 adults with PCD had registered within the *Living with PCD* study, 265 of them (55%) completed the fertility questionnaire (Figure 1, Table 1). Most of them were women (180; 68%) with a median age of 44 years (IQR 33-54) at survey. The respondents came from 33 countries with the largest number from the United States (41; 15%), England (39; 15%), Germany (37; 14%) and Switzerland (26; 10%). Compared with non-responders, people who completed the fertility questionnaire were older (Supplementary Table S3).

**Table 1:** Characteristics of adults with primary ciliary dyskinesia (PCD) participating in *Living with PCD* who completed the fertility questionnaire, overall and by sex (N = 265).

|  |  | Overall | Men | Women |
| --- | --- | --- | --- | --- |
|  |  | N (%)<br>N = 265 | n (%)<br>N = 85 | n (%)<br>N = 180 |
| **Age at survey** | Median (IQR; range) | 44 (33-54;<br>19-87) | 48 (34-56;<br>19-77) | 43 (33-52;<br>19-87) |
|  | 18-30 y | 47 (18) | 12 (14) | 35 (19) |
|  | 31-45 y | 100 (38) | 28 (33) | 72 (40) |
|  | > 45 y | 118 (45) | 45 (53) | 73 (41) |
| **Fertility information** | Tried to conceive | 168 (63) | 50 (59) | 118 (66) |
|  | Not (yet) tried to conceive | 87 (33) | 30 (35) | 57 (32) |
|  | Tried to conceive for less than 12 months | 10 (4) | 5 (6) | 5 (3) |
|  | **Available fertility status** | **n = 168** | **n = 50** | **n = 118** |
|  | Fertile | 57 (34) | 11 (22) | 46 (39) |
|  | Infertile | 111 (66) | 39 (78) | 72 (61) |
|  | Achieved pregnancy | 115 (68) | 28 (56) | 87 (74) |
|  | Did not achieve pregnancy | 53 (32) | 22 (44) | 31 (26) |
| **Countries<sup>a</sup>** | United States | 41 (15) | 12 (14) | 29 (16) |
|  | England | 39 (15) | 11 (13) | 28 (16) |
|  | Germany | 37 (14) | 9 (11) | 28 (16) |
|  | Switzerland | 26 (10) | 9 (11) | 17 (9) |
|  | Italy | 19 (7) | 8 (9) | 11 (6) |
|  | France | 16 (6) | 6 (7) | 10 (6) |
|  | Australia | 12 (5) | 4 (5) | 8 (4) |
|  | Canada | 10 (4) | 1 (1) | 9 (5) |
|  | Netherlands | 9 (3) | 4 (5) | 5 (3) |
|  | Scotland | 7 (3) | 3 (4) | 4 (2) |
|  | Norway | 6 (2) | 3 (4) | 3 (2) |
|  | Denmark | 5 (2) | 2 (2) | 3 (2) |
|  | other European countries | 30 (11) | 12 (14) | 18 (10) |
|  | other countries | 8 (3) | 1 (1) | 7 (4) |

Abbreviations: IQR, interquartile range. PCD, primary ciliary dyskinesia. y, years. All characteristics are presented as n and column % unless otherwise stated. <sup>a</sup>Countries with N≥5 displayed in table, countries with N<5 were categorised into other European countries and other countries. Other European countries include Austria, Belgium, Croatia, Cyprus, Czech Republic, Hungary, Ireland, Jersey, Northern Ireland, Poland, Portugal, Spain, Sweden, and Wales. Other countries include Brazil, Cameroon, Hong Kong, Israel, Panama, Puerto Rico, and South Africa.

### Fertility and pregnancy outcomes of men

Among 50 men who had tried to conceive, 11 men (22%) were fertile, and 39 (78%) were infertile (Figure 2, Table 1). Of the infertile men, 28 had tried MAR, and 17 of them (61%) fathered a child with the help of MAR. Overall, 28 men (56%) fathered a child with or without the help of MAR. The men in our study were a median of 31 years old (IQR 28-35) when they first tried to conceive a child. Among 62 men who reported sperm analysis, only 3 (5%) had a normal test result (Supplementary Table S4).

**Figure 2:**
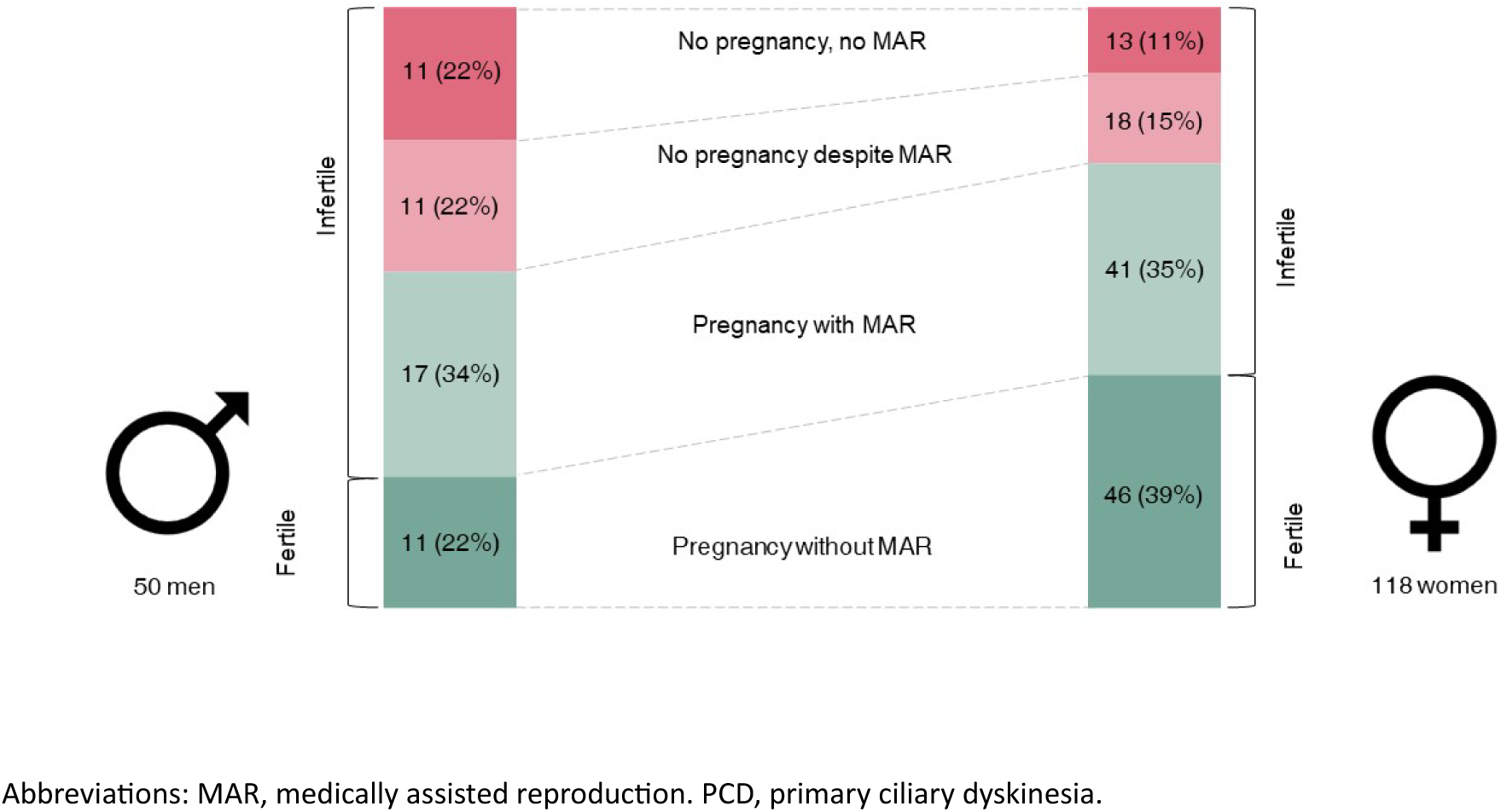
Fertility status of adults participating in *Living with PCD* who completed the fertility questionnaire and have ever tried to conceive, by sex (N = 168).

Overall, the 28 men who conceived reported a total of 50 pregnancies. In more than half of those pregnancies (28; 56%), they had used MAR. Types of fertilization most commonly used for successful pregnancies were ICSI (15; 41%), and IVF (9; 24%). Most of the pregnancies resulted in live births (39; 78%), but there were also 10 pregnancy losses or stillbirths (20%) reported by 7 men.

### Fertility and pregnancy outcomes of women

Among the 118 women who reported pregnancy attempts, 46 (39%) became pregnant naturally, and 72 women (61%) experienced infertility (Figure 2, Table 1). Among infertile women, 59 had used MAR, and 41 of them (69%) became pregnant with the help of MAR. A total of 87 women (74%) became pregnant at least once. The women in our study were a median of 30 years old (IQR 27-32) when they first tried to conceive a child (Supplementary Table S5).

Overall, 87 women who managed to become pregnant reported a total of 197 pregnancies (Table 2). Two-thirds (125; 63%) of the pregnancies were conceived without MAR. For successful pregnancies, IVF (55; 49%), ovarian stimulation with or without intra uterine insemination (26; 23%) or ICSI (20; 18%) were the most commonly used types of MAR. The 197 pregnancies resulted in 124 (63%) live births, 44 (22%) miscarriages, 15 (8%) ectopic pregnancies, and 11 (6%) abortions. 3 (2%) pregnancies were ongoing at the time of the study. Overall, 1 in 10 women who became pregnant had at least one ectopic pregnancy.

**Table 2:** Mode of fertilization and pregnancy outcomes for pregnancies reported by adults in *Living with PCD* who completed the fertility questionnaire, by sex (N = 247).

|  | All pregnancies<br>reported by<br>men | All pregnancies<br>reported by<br>women |
| --- | --- | --- |
|  | n (%)<br>N = 50 | n (%)<br>N = 197 |
| **Mode of fertilization** |  |  |
| Pregnancy without MAR | 22 (44) | 125 (63) |
| Pregnancy with MAR | 28 (56) | 70 (36) |
| Unknown mode of fertilization | 0 (0) | 2 (1) |
| Types of MAR used<sup>a</sup> | n = 37 | n = 113 |
| Ovarian stimulation | 0 (0) | 26 (23) |
| Artificial insemination | 2 (5) | 5 (4) |
| IVF | 9 (24) | 55 (49) |
| ICSI | 15 (41) | 20 (18) |
| Sperm donation | 4 (11) | 4 (4) |
| Oocyte donation | 0 (0) | 1 (1) |
| Partner used MAR | 7 (19) | 1 (1) |
| Unknown type of MAR | 0 (0) | 1 (1) |
| **Pregnancy outcomes** |  |  |
| Live birth | 39 (78)<sup>b</sup> | 124 (63)<sup>c</sup> |
| Pregnancy loss/stillbirth | 10 (20) | 44 (22) |
| Ectopic pregnancy | 0 (0) | 15 (8) |
| Abortion | 1 (2) | 11 (6) |
| Ongoing pregnancy | 0 (0) | 3 (2)<sup>d</sup> |

Abbreviations: ICSI, intracytoplasmic sperm injection. IVF, *in vitro* fertilization. MAR, medically assisted reproduction. PCD, primary ciliary dyskinesia. All characteristics are presented as n and column %. <sup>a</sup>Types of MAR exceed the number of pregnancies with MAR as more than one type of MAR was reported for each pregnancy on average. <sup>b</sup>Four live births were the result of sperm donations. <sup>c</sup>Five women reported the birth of twins (= 129 babies born). <sup>d</sup>One ongoing pregnancy was the result of an oocyte donation.

We estimated the prevalence of ectopic pregnancies among women to be 7.6 per 100 pregnancies (95% CI 4.7-12.2) and 12.1 per 100 live births (95% CI 7.5-19.0) (Table 3). Assuming that there were no other ectopic pregnancies among women who did not complete the questionnaire and that the number of pregnancies per woman was the same in the whole population, we calculated a minimum prevalence of ectopic pregnancies for the whole *Living with PCD* cohort of 4.2 per 100 pregnancies (95% CI 2.6-6.9) and 6.8 per 100 live births (95% CI 4.1-10.9).

**Table 3:** Frequency of ectopic pregnancies among all women participating in *Living with PCD* who completed the fertility questionnaire and among all women in *Living with PCD*.

| Ectopic pregnancies | Women in *Living with PCD* who completed fertility questionnaire | Minimum prevalence: All women in *Living with PCD*<sup>a</sup> |
| --- | --- | --- |
| - per 100 pregnancies | 7.6 (95% CI: 4.7-12.2) | 4.2 (95% CI: 2.6-6.9) |
| - per 100 live births | 12.1 (95% CI: 7.5-19.0) | 6.8 (95% CI: 4.1-10.9) |

Abbreviation: CI, confidence interval. PCD, primary ciliary dyskinesia. We calculated Wilson 95% confidence intervals for proportions. <sup>a</sup>We based the calculation of the minimum prevalence on two assumptions: women who did not complete the questionnaire had no ectopic pregnancy and all women in *Living with PCD* had the same median number of pregnancies. 180 women who completed the survey reported 197 pregnancies and 124 live births, 142 women who did not complete the survey would have 156 additional pregnancies and 98 additional live births. Overall, we would count 353 pregnancies and 222 live births among all women in *Living with PCD*. 15 ectopic pregnancies per 353 pregnancies of all women in *Living with PCD* = 4.2 ectopic pregnancies per 100 pregnancies. 15 ectopic pregnancies per 222 live births of all women in *Living with PCD* = 6.8 ectopic pregnancies per 100 live births.

Most of the nine women who reported ectopic pregnancies did not report any risk factors for their first ectopic pregnancy other than PCD, such as MAR use, or previous tubal surgery (Supplementary Table S6). For six women, the ectopic pregnancy was their first pregnancy. Five women reported more than one ectopic pregnancy and seven women reported one or more live births before or after the ectopic pregnancy.

### Association of fertility status with genotype

We assessed differences in fertility between genotypes for 46 people (11 men, 35 women) with available genetic and fertility information (Table 4). Infertility was reported in all individuals in the dynein assembly group [DNAAF3 n=1, ZMYND10 n=2, PIH1D3 (DNAAF6) n=1], the microtubular stabilization/nexin-dynein regulatory complex group [CCDC39 n=1, CCDC40 n=6, CCDC65 (DRC2) n=1] and the group with mutations in radial spoke and central complex (RSPH9 n=3, HYDIN n=1, RSPH1 n=1). All five women with a mutation in DNAH11 reported to be fertile. The pattern was more heterogeneous for other genes. Among women with a mutation in DNAH5, eight reported being fertile and seven being infertile.

**Table 4:** Gene groups and single PCD genes of men and women participating in *Living with PCD* with available genetic and fertility information who completed the fertility questionnaire, by fertility status (N = 46).

| Gene groups<sup>a</sup> | PCD gene | Total | Men |  | Women |  |
| --- | --- | --- | --- | --- | --- | --- |
|  |  |  | Fertile | Infertile | Fertile | Infertile |
|  |  | N = 46 | n = 3 | n = 8 | n = 16 | n = 19 |
| Dynein structure | DNAH5 | 17 | 2 | - | 8 | 7 |
|  | DNAH11 | 5 | - | - | 5 | - |
|  | DNAI1 | 4 | - | 1 | 3 | - |
|  | DNAH9 | 1 | - | 1 | - | - |
|  | ODAD1<br>(CCDC114) | 1 | - | 1 | - | - |
| Dynein assembly | DNAAF3 | 1 | - | - | - | 1 |
|  | ZMYND10 | 2 | - | - | - | 2 |
|  | PIH1D3<br>(DNAAF6) | 1 | - | 1 | - | - |
| Microtubular<br>stabilization/nexin-dynein regulatory<br>complex | CCDC39 | 1 | - | 1 | - | - |
|  | CCDC40 | 6 | - | 1 | - | 5 |
|  | CCDC65<br>(DRC2) | 1 | - | 1 | - | - |
| Radial spoke and<br>central complex | RSPH9 | 3 | - | - | - | 3 |
|  | HYDIN | 1 | - | - | - | 1 |
|  | RSPH1 | 1 | - | 1 | - | - |
| Other function | RPGR | 1 | 1 | - | - | - |

Abbreviation: PCD, primary ciliary dyskinesia. <sup>a</sup>We grouped affected genes according to the corresponding ultrastructural defect, as done previously (Shoemark, et al., 2021).

## Discussion

In the largest study on fertility in PCD to date, we showed a high prevalence of infertility among people with PCD, with two-thirds of women and four-fifths of men reporting difficulties conceiving. Notably, two-thirds of infertile individuals who underwent MAR successfully conceived. We counted 15 ectopic pregnancies in our study which corresponds to a prevalence of ectopic pregnancies of around 8 ectopic pregnancies per 100 pregnancies.

### Strengths and limitations

For the first time in PCD, we collected data on fertility status from adults with PCD worldwide. By directly asking people with PCD, we not only collected information on live births but also on other birth outcomes that are often not well recorded in medical charts to ensure accurate prevalence estimates. However, we were not able to validate clinical data obtained from participant self-reports due to the anonymous study design, which is likely to lead to recall bias. For example, we cannot exclude that miscarriages of unknown localisation have been be incorrectly described by patients as ectopic pregnancies in some instances. People struggling with fertility might have been more likely to complete the fertility questionnaire, which could have led to selection bias and influenced our prevalence estimates. Calculating the minimum prevalence of ectopic pregnancies for the entire study population confirmed the robustness of our findings. However, it is still possible that women who did not complete the questionnaire had more pregnancies. Due to the complexity of fertility status, which depends on various factors, we cannot causally link infertility to PCD in our study population. Other reasons for infertility include partner’s fertility problems, infrequent sexual intercourse, sexually transmitted diseases, or other urological or gynaecological conditions, some of which we have assessed in our questionnaire (Supplementary Tables S4 & S5). Without a clinical assessment, we cannot determine the definite reason for infertility.

### Comparison with previous research and general population

Our findings quantified the high risk of infertility among individuals with PCD confirming differences of fertility status between men and women. Our infertility prevalence estimates were higher (66% overall) than those estimated in the systematic review and meta-analysis from 2016 where infertility in seven studies was ranging from 15 to 79% with a weighted mean of 30% (Goutaki, et al., 2016). In three studies with results stratified by sex, the authors estimated male infertility at 100%, but most studies included only few male adults. In our study, 22% of the men fathered a child without the help of MAR. Our results are in line with results from the largest study on infertility to date by Vanaken from 2017. They assessed fertility in a cohort of patients recruited from PCD hospitals in France and Belgium and estimated the risk of infertility among 36 women to be 61% and among 49 men to be 76% (Vanaken, et al., 2017). In a narrative review published in 2023, prevalence was not estimated as the quality of evidence from case reports was not considered good enough (Newman, et al., 2023).

We showed an increased risk of ectopic pregnancies among women with PCD compared with the general population. Limited data on ectopic pregnancies among women in the general population of high-income countries estimates the risk to be at maximum 2 ectopic pregnancies per 100 pregnancies (Jurkovic and Wilkinson, 2011, Panelli et al., 2015, Van Den Eeden et al., 2005). Our results suggest that in women with PCD, this risk is 3.8-fold (95% CI 2.4-6.1) increased. To date, data on ectopic pregnancies among women with PCD is controversial. Both, spontaneous pregnancies and infertility have been reported for women with dyskinetic ciliary activity (Bleau et al., 1978, Lurie et al., 1989, Marchini, et al., 1992, McComb et al., 1986, Raidt, et al., 2015). Even though, pathophysiological considerations suggest an increased risk of ectopic pregnancy, the involvement of cilia in the transport of the oocyte in the uterine tractus and, consequently, female infertility is not well understood. Only four reports of ectopic pregnancies among women with PCD existed before this study (Blyth and Wellesley, 2008, Lin, et al., 1998, McLean and Claman, 2000, Newman, et al., 2023) with Vanaken not reporting any ectopic pregnancies among their 36 women (Vanaken, et al., 2017). We will need large clinical studies to confirm our results.

We provided new insights into the associations of fertility with genotype. In line with previous research, we showed that people with mutations in CCDC40 are more likely to be infertile (Liu et al., 2021, Vanaken, et al., 2017). In contrast, DNAH5 has not previously been associated with infertility in women (Newman, et al., 2023), but 7 out of 15 women in our study with a mutation in DNAH5 were infertile. Conversely, for DNAH11 we reported only fertile individuals, whereas previous publications showed both fertile and infertile individuals (Brennan et al., 2021, Newman, et al., 2023). We have also confirmed the importance of assessing other factors that affect fertility when interpreting the results. For example, a man with a CCDC114 mutation, which is not associated with a higher risk of infertility (Kos et al., 2022), was found to be infertile, possibly due to another severe condition he reported.

### Interpretation and implication for future research

Fertility is a topic that needs to be addressed in regular PCD care and people with PCD need to be informed about their possible increased risk of infertility (Schreck et al., 2024). However, not every person with PCD is infertile, which needs to be communicated as well. Additionally, we underlined the usefulness and effectiveness of MAR in PCD and suggest that it should be offered to infertile patients with PCD early on. For people with PCD who do not wish to get pregnant, adequate birth control should be in place. Women who are trying to conceive should be made aware of the potentially increased risk of ectopic pregnancy so that they react early and consult their gynaecologist to confirm the intrauterine position of the pregnancy before symptoms arise. It is important to note that the risk of ectopic pregnancy in our population was not related to the use of MAR. Future studies will need to assess infertility among people with PCD in even larger clinical cohorts and study the efficacy of MAR in more detail.

## Conclusion

We quantified a high risk of infertility among people with PCD. MAR was often used successfully in PCD-related infertility. Women attempting conception should be aware of the potentially elevated risk of ectopic pregnancies and seek systematic early consultation to confirm intrauterine pregnancy (4-5 weeks of gestation). Fertility, efficacy of MAR and risk for adverse pregnancy outcomes differ between people with PCD—depending on genotypes—, and close monitoring and support might be needed from fertility specialists to increase chances of successful conception.

## Authors’ roles

LD Schreck, ESL Pedersen, M Goutaki and CE Kuehni made substantial contributions to the study design. LD Schreck, ESL Pedersen, K Dexter, M Manion, N Massin, B Maitre, M Goutaki and Claudia E Kuehni made substantial contributions to the study concept and were involved in the design of the fertility questionnaire. K Dexter, M Manion, N Massin, B Maitre made substantial contribution to the interpretation of the data. LD Schreck analysed data and drafted the manuscript. LD Schreck, ESL Pedersen, K Dexter, M Manion, N Massin, B Maitre, M Goutaki, and CE Kuehni critically revised and approved the manuscript.

## Acknowledgements

We thank all participants and their families, and we thank the PCD support groups and physicians who advertised the study. We thank our collaborators who helped set up the *Living with PCD* study: Cristina Ardura, Yin Ting Lam, Christina Mallet, Helena Koppe, Dominique Rubi from the University of Bern and Jane S Lucas and Amanda Harris from the University Hospital Southampton. We thank Sophie Christin-Maitre and Lara Gonçalves Pissini for their help in the design of the fertility questionnaire.

## Funding

Our research was funded by the Swiss National Science Foundation, Switzerland (SNSF 320030B_192804/1), the Swiss Lung Association, Switzerland (2021-08_Pedersen), and we also received support from the PCD Foundation, United States; the Verein Kartagener Syndrom und Primäre Ciliäre Dyskinesie, Germany; PCD Support UK, United Kingdom; and PCD Australia, Australia. M. Goutaki received funding from the Swiss National Science Foundation, Switzerland (PZ00P3_185923). B. Maitre participates in the RaDiCo-DCP funded by INSERM France. Study authors participate in the BEAT-PCD Clinical Research Collaboration supported by the European Respiratory Society.

## Conflict of interest

All authors declare no conflict of interest.

## Data availability statement

*Living with PCD* data is available upon reasonable request by contacting Claudia Kuehni.

---

**Supplementary Table S1:** Formulation of relevant questions from the English adult baseline questionnaire and the English female and male fertility questionnaire of the *Living with PCD* study.

| Question | Answer category |
| --- | --- |
| **Baseline Questionnaire** |  |
| What is your sex? | Male; Female; Other |
| Which year were you born? | *text (integer)* |
| Which country do you live in? | List of all countries worldwide |
| Which other country? | *text* |
| What is your height in centimeter? If you would rather enter feet and inches, please go to the next question | *text (integer)* |
| What is your height in feet and inches? | *text* |
| What is your weight in kilogram? If you would rather enter weight in pounds, please go to the next question | *text (integer)* |
| What is your weight in pounds? | *text (integer)* |
| Do you smoke? | No, I have never smoked; Yes, rarely; Yes, daily; Ex-smoker, I don't smoke anymore |
| Are any of your organs in a different position compared to most people? (E.g. the heart on the right side instead of on the left) | No; Yes; I don't know |
| Which year were you diagnosed with PCD? | *text (integer)* |
| How old were you, when you were diagnosed with PCD? | *text (integer)* |
| Have you had diagnostic tests for PCD? | No; Yes |
| Have you had a genetic test (looking for genes that cause PCD)? | No; Yes; I don't know/I cannot remember |
| Were any genes found that cause PCD? | No; Yes; I don't know/ I cannot remember/ waiting for results |
| Which gene was found? | List of all known PCD genes (in 2020) |
| Which gene was found? (If a second gene was found) | List of all known PCD genes (in 2020) |
| **Fertility questionnaire** |  |
| Have you ever seen a fertility expert (= a doctor who is specialized in fertility)? | No;<br>No, but I have an appointment / I am waiting to be referred;<br>No, but I would like to;<br>Yes |
| How old were you when you first saw a fertility expert? If you cannot remember the exact age, please give an approximate age. | *text (integer)* |
| Who referred you to the fertility expert? | I was referred by my PCD physician.;<br>I was referred by a doctor other than my PCD physician (e.g. my general practitioner).;<br>I had to ask for a referral.;<br>I had to organize it myself.;<br>Other |
| Please specify who referred you to the fertility expert | *text* |
| Have you ever been told by a health care professional that you may have problems with your fertility or that you are infertile? | No; Yes; I don't know |
| What reasons for your fertility problems were mentioned? (Tick all that apply) | Reason related to PCD;<br>Reason related to other causes;<br>Unknown cause of fertility problems at that time, PCD only diagnosed after;<br>Unknown reason;<br>I don't know |
| Please specify what reason for your fertility problems was mentioned | *text* |
| [only for females] How regular is/was your period during your reproductive years between 18 and 40 years (when you are/were not using hormonal contraceptives, e.g. the pill)? | My period is/was usually regular, occurring every 25-35 days.;<br>My period is/was sometimes regular, sometimes irregular, but my menstrual cycle is only a few days longer or shorter than 25-35 days.;<br>My period is/was totally irregular or absent.;<br>Other;<br>I don't know / I don't remember |
| [only for females] Please specify how your period is/was during your reproductive years | *text* |
| [only for females] Have you been diagnosed with any of the following conditions that affect fertility which are not known to be related to PCD? (Tick all that apply) | No;<br>Polycystic ovaries (many fluid-filled sacs in the ovaries);<br>Endometriosis (condition where tissue similar to the tissue in the uterus starts to grow in other places of the body);<br>Other problems |
| [only for females] Please specify which condition you have been diagnosed with | *text* |
| Have you ever had any tests to check your fertility? | No; Yes |
| What tests did you have to check your fertility? | [only for females] Checking the flow through the uterine tubes (tubal patency testing, HyCoSy). A liquid is injected into the uterus and then checked by ultrasound or radiography to see if it reaches the abdomen.;<br>Measurement of fertility hormones;<br>Other |
| Please specify what other tests were performed | *text* |
| How old were you when the tests were performed? If you cannot remember the exact age, please give an approximate age. | *text (integer)* |
| [only for females] What was the result of the tubal patency testing? | Both tubes were open/permeable.;<br>One tube was blocked.;<br>Both tubes were blocked.;<br>I don't know |
| Were the levels of fertility hormones normal? | No; Yes; I don't know |
| [only for females] Did you have to take medication or have you had any surgical procedures on your ovaries or uterus? | No;<br>Yes, I had to take medication to have regular ovulation.;<br>Yes, I had surgical procedures on my ovaries or uterus.;<br>Yes, other |
| [only for females] Please specify what treatment or surgical procedure you had | text |
| *For the following questions, only the formulation for females is reported* |  |
| Have you ever tried to become pregnant? | No;<br>Yes, but it did not work / we are still trying;<br>Yes, it worked |
| Have you ever been pregnant? This also includes pregnancies that were terminated or resulted in miscarriages. | No; Yes, once; Yes, twice; Yes, three times;<br>Yes, four times; Yes, five times; Yes, more than five times |
| How many times have you been pregnant? This also includes pregnancies that were terminated or resulted in miscarriages. | Once; Twice; Three times; Four times; Five times; More than five times |
| How old were you when you were pregnant the first time? | text (integer) |
| Was the first/second/third/fourth/fifth/sixth pregnancy a natural pregnancy or a pregnancy as a result of fertility treatment? | Natural pregnancy; Pregnancy with help of fertility treatment; I don't know |
| How long had you been trying to conceive for the first/second/third/fourth/fifth/sixth pregnancy? | < 6 months; 6 to 12 months; 1 to 2 years;<br>More than 2 years; I don't know / I don't remember |
| What fertility therapy was used for the first/second/third/fourth/fifth/sixth pregnancy? (Tick all that apply) | I received medication that stimulated my ovaries (ovarian stimulation).;<br>My partner's semen (sperm) was directly inserted into my uterus (artificial insemination).;<br>One of my eggs was combined with my partner's sperm outside the body (in vitro fertilization (IVF)).;<br>My partner's sperm was injected directly into an egg (intracytoplasmic sperm injection (ICSI)).;<br>We used donated sperm.;<br>An egg was donated to me (oocyte donation).;<br>My partner had to undergo fertility treatment.;<br>Other;<br>I don't know |
| Please specify what fertility therapy was used for the first/second/third/fourth/fifth/sixth pregnancy | text |
| What was the outcome of the first/second/third/fourth/fifth/sixth pregnancy? | The baby was born alive.;<br>The pregnancy was outside my uterus and had to be terminated (ectopic pregnancy).;<br>The pregnancy was ended with an intervention (abortion).; |
|  | The baby was lost during pregnancy or birth (miscarriage or stillbirth).;<br>Other |
| Please specify the outcome of the first/second/third/fourth/fifth/sixth pregnancy | *text* |
| [only for females] Did your lung health get worse during the first pregnancy? (Tick all that apply) | No;<br>Yes, I had new or worsening symptoms related to PCD.;<br>Yes, I saw my PCD doctor more frequently.;<br>Yes, I had to change or increase my medication during pregnancy.;<br>Yes, I was admitted to the hospital for PCD-related issues. |
| How old were you when you tried getting pregnant the first time? | *text (integer)* |
| How long have you been trying to get pregnant? | < 6 months; 6 to 12 months; 1 to 2 years;<br>More than 2 years; I don't know |
| Do you know if the reasons for not conceiving a child are related to your fertility problems or your partner's? (Tick all that apply) | My fertility problems; My partner's fertility problems; Unknown or unexplained fertility problems; I don't know |
| Did you use any fertility therapy? | No; Yes |
| Why did you not use any fertility therapy? (Tick all that apply) | It was not available where I lived.; It was not covered by the health insurance.; I could not afford it.; I did not want to.; Other; I don't know |
| Please specify the reason for not using fertility therapy | *text* |
| What fertility therapy was used? (Tick all that apply) | I received medication that stimulated my ovaries (ovarian stimulation).;<br>My partner's semen (sperm) was directly inserted into my uterus (artificial insemination).;<br>One of my eggs was combined with my partner's sperm outside the body (in vitro fertilization (IVF)).;<br>My partner's sperm was injected directly into an egg (intracytoplasmic sperm injection (ICSI)).;<br>We used donated sperm.;<br>An egg was donated to me (oocyte donation).;<br>My partner had to undergo fertility treatment.;<br>Other;<br>I don't know |
| Please specify what fertility therapy was used | *text* |

**Supplementary Table S2:** Definition of abnormal fertility tests and other factors associated with fertility assessed in the *Living with PCD* study.

| Test/Condition | Definition | Sex |
| --- | --- | --- |
| **BMI > 30/≤ 18** | Adults who reported a BMI > 30 or ≤ 18 in the baseline questionnaire.<br>To calculate body mass index (BMI), we used self-reported height and weight data from the *Living with PCD* baseline questionnaire. We calculated BMI by dividing weight in kilograms by height in meters squared (kg/m<sup>2</sup>). | Both |
| **Ever smoked/smoking** | Adults who reported that they are currently smoking or that they are ex-smokers in the baseline questionnaire. | Both |
| **Abnormal levels of fertility hormones** | Adults who reported to have abnormal levels of fertility hormones. | Both |
| **Other condition** | Adults who reported having any other condition that could affect fertility in the fertility questionnaire:<br>- for women: removal of an ovary, low egg count and quality, ovarian dermoid cysts, borderline tumors of the ovary, uterine fibroids, malformation of the uterus, papillomavirus infection.<br>- for men: varicocele. | Both |
| **Period totally irregular or absent** | Women who reported that their period was totally irregular or absent. | Women |
| **Endometriosis** | Women who reported suffering from endometriosis. | Women |
| **Polycystic ovaries** | Women who reported suffering from polycystic ovaries. | Women |
| **Problems with tubal patency** | Women who reported that either one or both fallopian tubes were blocked. | Women |
| **Abnormal ultrasound of testicles** | Men who reported abnormal ultrasound of testicles. | Men |
| **Abnormal result of sperm analysis** | Men who reported an abnormal sperm analysis such as low sperm concentration, low sperm mobility, low sperm vitality, too few sperm with normal shape or other reasons for abnormal sperm result. | Men |

Abbreviations: BMI, body mass index. PCD, primary ciliary dyskinesia.

**Supplementary Table S3:** Comparison of basic characteristics of adults participating in *Living with PCD* who did or did not complete the questionnaire on fertility (N = 482).

|  |  | Completed<br>questionnaire | Did not complete<br>questionnaire |
| --- | --- | --- | --- |
|  |  | n (%)<br>n = 265 | n (%)<br>n = 217 |
| **Age at survey** | Median (IQR) | 44 (33-54) | 36 (26-47) |
|  | 18-30 y | 47 (37) | 80 (63) |
|  | 31-45 y | 100 (56) | 78 (44) |
|  | > 45 y | 118 (67) | 59 (33) |
| **Sex** | Women | 180 (56) | 142 (44) |
|  | Men | 85 (54) | 73 (46) |
|  | Other | 0 (0) | 2 (100) |
| **Countries<sup>a</sup>** | United States | 41 (47) | 47 (53) |
|  | England | 39 (46) | 45 (54) |
|  | Germany | 37 (65) | 20 (35) |
|  | Switzerland | 26 (76) | 8 (24) |
|  | Italy | 19 (50) | 19 (50) |
|  | France | 16 (59) | 11 (41) |
|  | Australia | 12 (71) | 5 (29) |
|  | Canada | 10 (71) | 4 (29) |
|  | Netherlands | 9 (60) | 6 (40) |
|  | Scotland | 7 (78) | 2 (22) |
|  | Norway | 6 (67) | 3 (33) |
|  | Denmark | 5 (56) | 4 (44) |
|  | Spain | 4 (44) | 5 (56) |
|  | Sweden | 3 (43) | 4 (57) |
|  | Austria | 3 (50) | 3 (50) |
|  | Ireland | 3 (60) | 2 (40) |
|  | other European countries | 17 (65) | 9 (35) |
|  | other countries | 8 (29) | 20 (71) |

Abbreviations: IQR, interquartile range. PCD, primary ciliary dyskinesia. y, years. All characteristics are presented as n and row % or as median and interquartile range. <sup>a</sup>Countries with N≥5 adult participants displayed in table, countries with N<5 were categorised into other European countries and other countries.

**Supplementary Table S4:** Information about fertility characteristics of men participating in *Living with PCD* who completed the fertility questionnaire, by fertility status (N = 85).

|  | All men | Fertile | Infertile | Unknown fertility status<sup>a</sup> |
| --- | --- | --- | --- | --- |
|  | n (%)<br>N = 85 | n (%)<br>n = 11 | n (%)<br>n = 39 | n (%)<br>n = 35 |
| **Age at first pregnancy attempt** | **n = 50** | **n = 11** | **n = 39** | **NA** |
| Median (IQR; range) | 31 (28-35; 19-45) | 29 (28-32, 19-38) | 31 (28-35; 20-45) | NA |
| 18-30 y | 23 (46) | 8 (73) | 15 (38) | NA |
| 31-45 y | 25 (50) | 3 (27) | 22 (56) | NA |
| Missing | 2 (4) | 0 (0) | 2 (5) | NA |
| **Sperm analysis done** | **n = 62** | **n = 6** | **n = 37** | **n = 19** |
| Normal sperm analysis | 3 (5) | 1 (17) | 1 (3) | 1 (5) |
| Low sperm concentration | 25 (40) | 2 (33) | 14 (38) | 9 (47) |
| Low sperm mobility | 47 (76) | 2 (33) | 32 (86) | 13 (68) |
| Low sperm vitality | 7 (11) | 0 (0) | 7 (19) | 0 (0) |
| Too few sperm with normal shape | 6 (10) | 1 (17) | 3 (8) | 2 (11) |
| Other reasons for abnormal sperm result | 2 (3) | 1 (17) | 0 (0) | 1 (5) |
| I don't know result | 1 (2) | 0 (0) | 0 (0) | 1 (5) |
| **Conditions that affect fertility** | **17 (20)** | **0 (0)** | **12 (31)** | **5 (14)** |
| - Ever smoked/smoking | 14 (16) | 0 (0) | 9 (23) | 5 (14) |
| - Abnormal ultrasound of testicles | 3 (4) | 0 (0) | 2 (5) | 1 (3) |
| - Abnormal levels of fertility hormones | 3 (4) | 0 (0) | 3 (8) | 0 (0) |
| - Other condition | 1 (1) | 0 (0) | 1 (3) | 0 (0) |

Abbreviations: BMI, body mass index. IQR, interquartile range. NA, not applicable. PCD, primary ciliary dyskinesia. y, years. All characteristics are presented as n and column % unless otherwise stated. <sup>a</sup>We classified people as unknown fertility status if they reported not having tried to conceive or if they tried for less than 12 months.

**Supplementary Table S5:** Information about fertility characteristics of women participating in *Living with PCD* who completed the fertility questionnaire, by fertility status (N = 180).

|  | All women | Fertile | Infertile | Unknown fertility status<sup>a</sup> |
| --- | --- | --- | --- | --- |
|  | n (%)<br>N = 180 | n (%)<br>n = 46 | n (%)<br>n = 72 | n (%)<br>n = 62 |
| **Age at first pregnancy attempt** | **n = 118** | **n = 46** | **n = 72** | **NA** |
| Median (IQR; range) | 30 (27-32; 16-40) | 28 (24-33; 16-39) | 30 (27-32; 19-40) | NA |
| 16-30 y | 75 (64) | 32 (70) | 43 (60) | NA |
| 31-45 y | 42 (36) | 14 (30) | 28 (39) | NA |
| Missing | 1 (1) | 0 (0) | 1 (1) | NA |
| **Other conditions associated with infertility** | **78 (43)** | **22 (48)** | **36 (50)** | **20 (32)** |
| - BMI > 30, ≤ 18 | 25 (14) | 9 (20) | 12 (17) | 4 (6) |
| - Ever smoked/smoking | 16 (9) | 4 (9) | 4 (6) | 8 (13) |
| - Period totally irregular/absent | 11 (6) | 2 (4) | 5 (7) | 4 (6) |
| - Endometriosis | 16 (9) | 3 (7) | 10 (14) | 3 (5) |
| - Polycystic ovaries | 12 (7) | 2 (4) | 8 (11) | 2 (3) |
| - Problems with tubal patency | 7 (4) | 2 (4) | 5 (7) | 0 (0) |
| - Abnormal levels of fertility hormones | 6 (3) | 1 (2) | 5 (7) | 0 (0) |
| - Other condition | 9 (5) | 2 (4) | 5 (7) | 2 (3) |

Abbreviations: BMI, body mass index. IQR, interquartile range. NA, not applicable. PCD, primary ciliary dyskinesia. y, years.
All characteristics are presented as n and column % unless otherwise stated. <sup>a</sup>We classified people as unknown fertility status if they reported not having tried to conceive or if they tried for less than 12 months.

**Supplementary Table S6:** Characteristics of women participating in *Living with PCD* who completed the fertility questionnaire and reported an ectopic pregnancy (n = 9).

|  | #1 | #2 | #3 | #4 | #5 | #6 | #7 | #8 | #9 |
| --- | --- | --- | --- | --- | --- | --- | --- | --- | --- |
| Age at first ectopic pregnancy (in y)<sup>a</sup> | 21-25<sup>b</sup> | 21-25<sup>b</sup> | 36-40 | 21-25 | 26-30 | 16-20 | 31-35 | 31-35 | 26-30<sup>b</sup> |
| BMI | 28 | 34 | 29 | 34 | 30 | 27 | 19 | 24 | 18 |
| Number of ectopic pregnancies | 2 | 3 | 1 | 1 | 2 | 2 | 1 | 2 | 1 |
| Mode of fertilization for ectopic pregnancies | both natural pregnancies | 2 natural pregnancies, 1 NA | natural pregnancy | natural pregnancy | both natural pregnancies | both natural pregnancies | IVF | both natural pregnancies | IVF |
| Pregnancy rank of ectopic pregnancy | first, second | first, second, fifth | third | first | first, second | first, second | first | second, third | second |
| Miscarriage before ectopic pregnancy | No | No | Yes | No | No | No | No | No | Yes |
| Number of live births (mode of fertilization) | 3 (all with ICSI) | 2 (both with IVF) | 1 (natural pregnancy) | 1 (natural pregnancy) | 1 (natural pregnancy) | 0 | 2 (both with IVF) | 2 (1 natural pregnancy, 1 with IVF) | 0 |
| Ever smoked/smoking | No | No | No | No | No | Yes | No | No | No |
| Tubal surgery | No | No | No | Yes | No | No | No | No | No |
| Tubal patency | Both open | Not tested | Both open | Not tested | Not tested | Not tested | Not tested | Not tested | Not tested |
| Endometriosis | No | No | No | No | No | No | No | No | Yes |
| Situs inversus | No | No | Yes | Yes | Yes | Yes | Yes | No | Yes |
| Gene | NA | NA | NA | DNAH5 | DNAH5 | NA | NA | NA | NA |

Abbreviation: BMI, body mass index. ICSI, intracytoplasmic sperm injection. IVF, *in vitro* fertilization. MAR, medically assisted reproduction. NA, not available. PCD, primary ciliary dyskinesia.
<sup>a</sup>To ensure anonymity, we only provide age groups. <sup>b</sup>Age at ectopic pregnancy was not reported. We used age at first pregnancy attempt (for pregnancies without MAR) or age at fertility specialist visit (for pregnancies with the use of MAR) as a proxy.

## Notes

### Competing Interest Statement

The authors have declared no competing interest.

### Author Declarations

The cantonal ethics committee of Bern, Switzerland, approved the study (study ID: 2020-00830).

